# High efficacy of face masks explained by characteristic regimes of airborne SARS-CoV-2 virus abundance

**DOI:** 10.1101/2020.09.10.20190348

**Authors:** Yafang Cheng, Nan Ma, Christian Witt, Steffen Rapp, Philipp Wild, Meinrat O. Andreae, Ulrich Pöschl, Hang Su

## Abstract

Airborne transmission by droplets and aerosols is important for the spread of viruses and face masks are a well-established preventive measure, but their effectiveness for mitigating COVID-19 is still under debate. We show that variations in mask efficacy can be explained by different regimes of virus abundance. For SARS-CoV-2, the virus load of infectious individuals can vary by orders of magnitude, but we find that most environments and contacts are in a virus-limited regime where simple surgical masks are highly effective on individual and population-average levels, whereas more advanced masks and other protective equipment are required in potentially virus-rich indoor environments such as medical centers and hospitals. Due to synergistic effects, masks are particularly effective in combination with other preventive measures like ventilation and distancing.

**One Sentence Summary:** Face masks are highly effective due to prevailing virus-limited environments in airborne transmission of COVID-19.

## Main Text

Airborne transmission is regarded as one of the main pathways for the transmission of viruses that lead to infectious respiratory deceases, including the severe acute respiratory syndrome coronavirus 2 (SARS-CoV-2) (*1*), and wearing face masks has been widely advocated to minimize transmission and protect people. Though commonly used, the effectiveness of masks is still under debate. Compared to N95/FFP2 or N99/FFP3 respirators with very low particle penetration rates (around ~1-5%), surgical and similar masks exhibit higher and more variable penetration rates (around ~30-70%) (*2, 3*). Given the large number of particles emitted upon respiration and especially upon sneezing or coughing (*4*), the number of respiratory particles that may penetrate masks is substantial, which is one of the main reasons leading to doubts about their efficacy in preventing infections. Moreover, randomized clinical trials show inconsistent results, with some studies reporting only a marginal benefit or no effect of mask use (*5*). Thus, surgical and similar masks are often considered to be ineffective. On the other hand, observational data show that regions or facilities with a higher percentage of the population wearing masks have better control of the coronavirus disease 2019 (COVID-19) (*6–8*). So how to explain the contrasting results and apparent inconsistency that masks with relatively high penetration rates may still have a significant impact on airborne virus transmission and the spread of COVID-19? Here, we combine knowledge and results of aerosol science and medical research with recent literature data to explain the contrasts and provide a basis for quantifying the efficacy of face masks.

When evaluating the effectiveness of masks, we want to understand and quantify its effect on the infection probability, *P*_inf_. Assuming that every single virus has the same chance to infect a person, *P*_inf_ can be calculated by

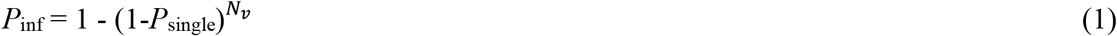

where *P*_single_ represents the infection probability of a single virus and *N*_v_ represents the total number of viruses to which the person is exposed (*9*). For airborne transmission, the infection probability *P*_inf_ for a given time period can be plotted as a function of inhaled virus number, *N*_v_.

Figure 1 illustrates the functional dependence of *P*_inf_ on *N*_v_ based on the exponential dose response model (Eq. 1) and scaled by the median infectious dose ID_v,50_ at which the probability of infection is 50% (*9*). It shows a highly nonlinear sensitivity of *P*_inf_ to changes of *N*_v_. Accordingly, the same percentage change of inhaled virus number may lead to different changes in *P*_inf_ depending on the absolute level of *N*_v_. In a virus-rich regime where *N*_v_ is much higher than ID_v,50_ (Figs. 1A and 1B), the probability of infection is close to unity and not sensitive to changes of *N*_v_. In this case, wearing a mask to reduce the inhaled amount by up to a factor of 10 may not suffice to prevent infection. In a virus-limited regime where *N*_v_ is close to or lower than ID_v,50_, however, *P*_inf_ strongly varies with *N*_v_, and reducing the inhaled number of airborne viruses by wearing a mask will lead to a significant reduction of the infection probability (Figs. 1C and 1D). Thus, we need to determine the regime of airborne virus abundance to understand the efficacy of wearing masks.

**Fig. 1.**
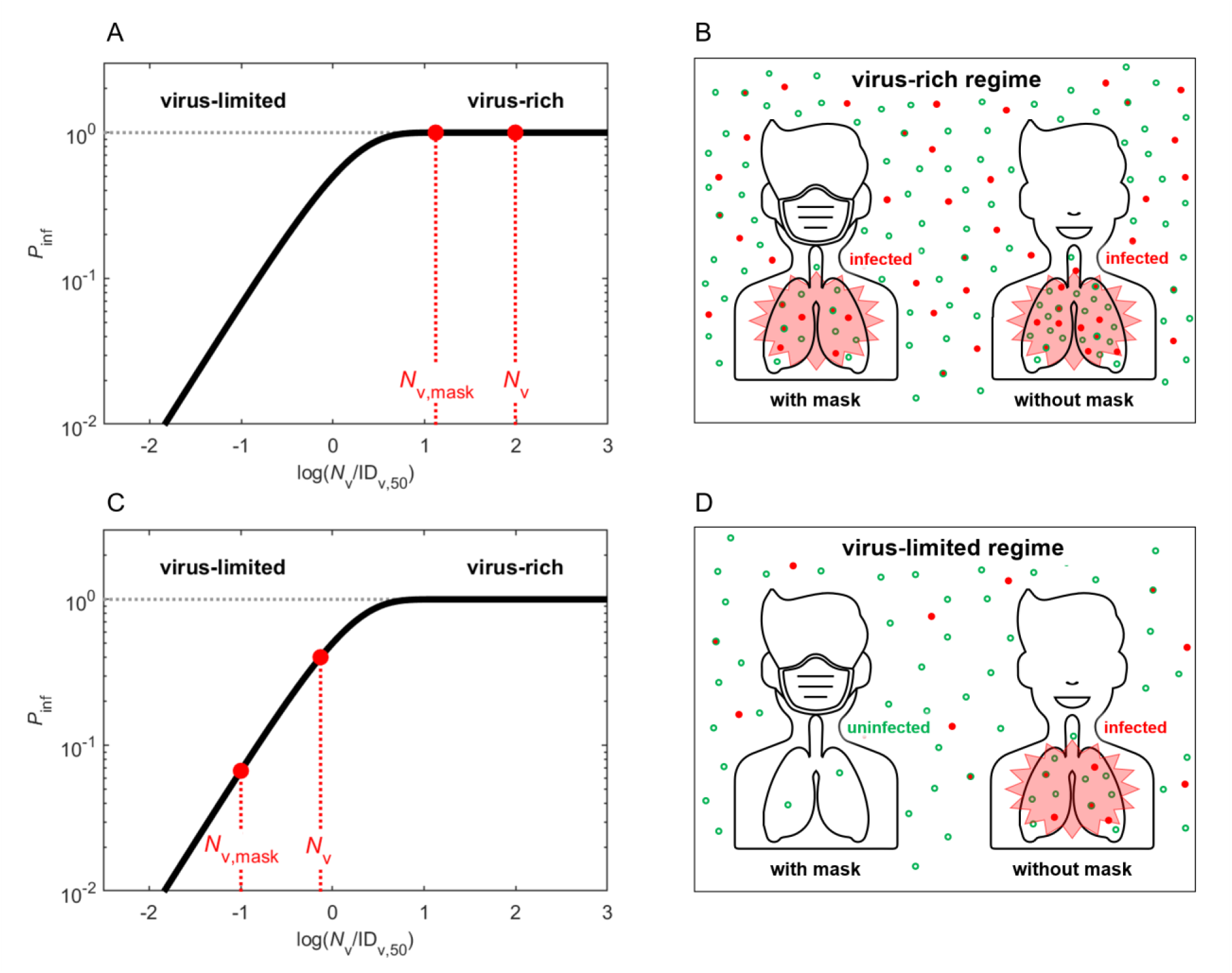
Schematic illustration of different regimes of abundance of respiratory particles and viruses. The solid curve represents the infection probability (*P*_inf_) as a function of inhaled virus number (*N*_v_) scaled by median infectious dose ID_v,50_ at which *P*_inf_ = 50%. In the virus-rich regime (**A, B**), the concentration of airborne viruses is so high, that both number of viruses inhaled with or without mask (*N*_v,mask_, *N*_v_) are much higher than ID_v,50_ and *P*_inf_ remains close to ~1 even if masks are used. In the virus-limited regime (**C, D**), *N*_v_ and *N*_v,mask_ are close to or lower than ID_v,50_ and *P*_inf_ decreases substantially when masks are used, even if the masks cannot prevent the inhalation of all respiratory particles. In panels B and D, the red dots represent viruses, and the open green circles represent respiratory particles.

Respiratory particles that may carry viruses are often used to visualize and represent the transmission of airborne viruses (*4*). Taking a representative average of respiratory activity (*10*), we find that a person typically emits a total number of about 3×10^6^ particles during a 30 min period (Sect S1). This very large number implies that indoor environments are usually in a respiratory particle-rich regime. Surgical masks with particle collection efficiencies around ~50% cannot prevent the release of millions of particles per person and their inhalation by others (green dots in Figs. 1B and 1D). In other words, the human-emitted particle number is so high that we cannot avoid inhaling particles generated by another person even when wearing a surgical mask. If every respiratory particle were to contain one or more viruses, indoor environments would often be in a virus-rich regime because the median infectious dose ID_v,50_ (virus number leading to 50% infection) for respiratory diseases is typically of the order of a few tens to thousands of viruses (*11–13*).

But does a respiratory particle-rich regime really imply a respiratory virus-rich regime? To answer this question, we investigate characteristic virus distributions in both exhaled air samples and indoor air samples. As we are not aware of any direct measurement of respiratory SARS-CoV-2 emission rates, we analyzed the results recently reported by Leung *et al*. (2020) (*10*) for a large number of samples of other types of viruses (n = 246; coronaviruses, influenza viruses, rhinoviruses) in respiratory particles with diameters < 5 *µ*m (“aerosol mode”) and > 5 *µ*m (“droplet mode”). As many samples in Leung *et al*. (2020) (*10*) returned a viral load signals below the detection limit, we reconstructed the mathematical expectation based on the percentage of positive cases and standard deviations (*σ*) of virus load distributions (detailed in Sect S2).

We find that usually just a minor fraction of exhaled respiratory particles contains viruses. In contrast to the high number of emitted respiratory particles, the number of viruses in 30-minute samples of exhaled air (*N*_v,30,ex_) are typically low with mean values around ~45 for coronaviruses (HCoV-NL63, -OC43, -229E and -HKU1), ~34 for influenza viruses (A and B), ~85 for rhinoviruses, and geometric standard deviations around ~1 (Fig. 2).

**Fig. 2.**
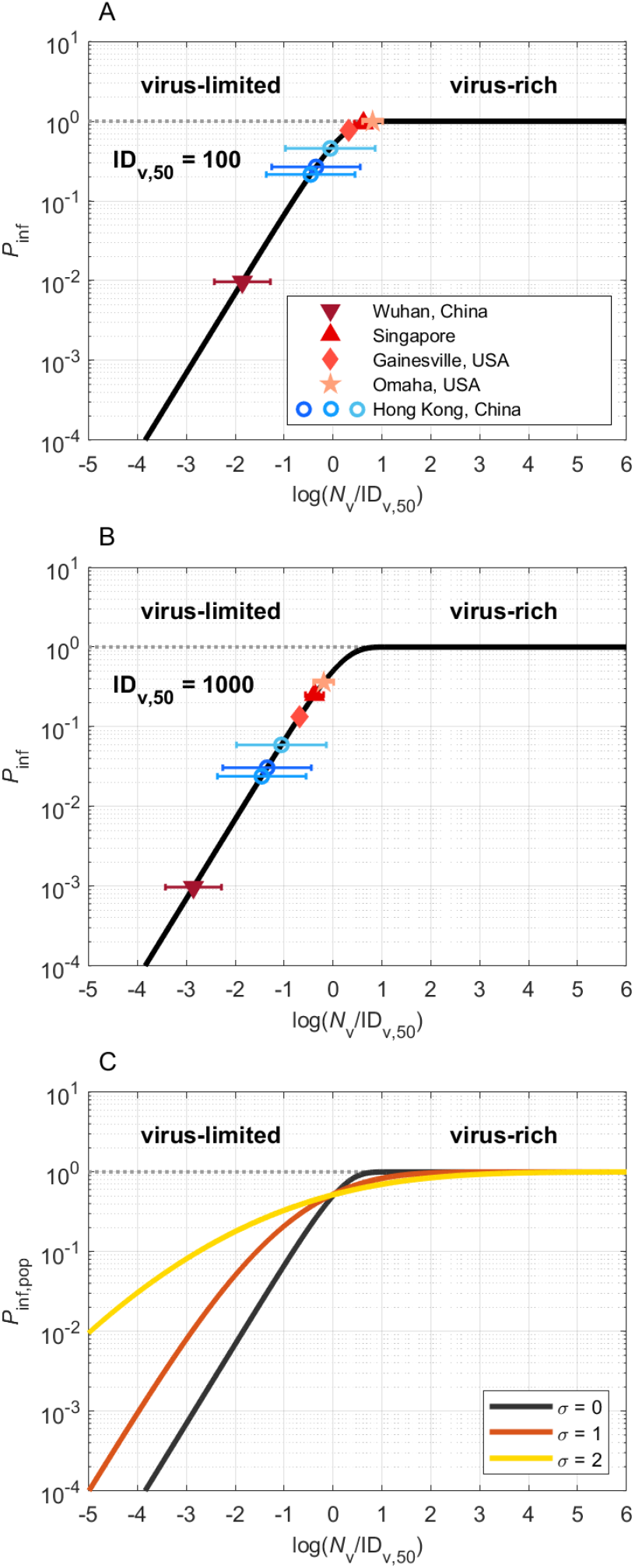
Infection probabilities and abundance regimes of SARS-CoV-2 and other respiratory viruses. **(A), (B)** Individual infection probabilities (*P*_inf_) plotted against inhaled virus number (*N*_v_) scaled by characteristic median infectious doses of ID_v,50_ = 100 or 1000, respectively. The colored data points represent the mean numbers of viruses inhaled during a 30-minute period in different medical centers in China, Singapore, and the USA, according to measurement data of exhaled coronavirus, influenza virus, and rhinovirus numbers (blue circles) and of airborne SARS-CoV-2 number concentrations (red symbols), respectively. The error bars are geometric standard deviations (blue circles). **(C)** Population-average infection probability (*P*_inf,pop_) curves assuming lognormal distributions of *N*_v_ with different standard deviations of *σ* = 0, 1, and 2, respectively.

The number of viruses exhaled by an infectious person, *N*_v,30,ex_, represents an upper limit for the number of viruses that can be inhaled by a susceptible person during a 30-minute contact with the infectious person. The blue data points (open circles) in Figs. 2A and 2B show the infection probabilities obtained by inserting the number of exhaled viruses (*N*_v,30,ex_) for the number of potentially inhaled viruses (*N*_v,30_) assuming median infectious doses of ID_v,50_ = 100 and 1000, respectively, which have been suggested as characteristic for SARS-CoV-2 and other respiratory viruses (*11–13*) (Sect S2). The resulting low values of log(*N*_v,30_/ ID_v,50_) clearly indicate a virus-limited regime.

For the SARS-CoV-2, few measurement data are available to date. In the Fangcang Hospital in Wuhan, China, treating mostly mild COVID-19 cases in a large and well-ventilated hall, the mean number concentration of airborne SARS-CoV-2 was 0.006 L^-1^ (*14*). By multiplication with an average breathing volume of 240 L during 30 minutes, we obtain a corresponding *N*_v,30_ value of ~1, which indicates a virus-limited regime and corresponds to *P*_inf_ values around ~1% or 0.1% for ID_v,50_ = 100 or 1000, respectively (Figs. 2A and 2B). For other medical centers in Singapore and the USA we obtained *N*_v,30_ mean values around ~200 to 600 with maximum values up to ~2000 (*15–17*), which are in a transition regime between virus-limited and virus-rich, corresponding to *P*_inf_ values around ~10-100% (Figs. 2A and 2B). In environments with such high virus number concentrations, surgical masks are not sufficient and more efficient masks are required to achieve substantial reductions of *P*_inf_. Under unfavorable conditions, e.g., at log(*N*_v,30_/ID_v,50_) ≈ log(2000/100) ≈ 1, even N95/FFP2 or N99/FFP3 masks are not sufficient to achieve low *P*_inf_ values, and further protective measures are required to prevent infections (e.g., hazmat suits, efficient ventilation, reduced occupation/circulation).

To check the consistency of our approach and results obtained with virus numbers measured in exhaled air and in ambient air, and for SARS-CoV-2 and other respiratory viruses, respectively, we designed a scenario emulating the numbers of patients, room size, and ventilation conditions reported for the Fangcang Hospital in Wuhan (Sect. S3). For this scenario, we calculated airborne virus concentrations based on the emission rates derived from Leung *et al*. (2020) for coronavirus, influenza viruses, and rhinoviruses. From these concentrations, we obtained *N*_v,30_ mean values in the range of ~0.08-0.2 which are of similar magnitude as the *N*_v,30_ value of ~1 derived from the measurements of airborne SARS-CoV-2 in the Fangcang Hospital (*14*). The consistency of these results and approaches are also supported by a recent comparison of viral load in respiratory tract fluids (*18*), where the loads observed for SARS-CoV-2 were similar to those of influenza B and rhinoviruses.

Different conditions of occupation, ventilation, circulation, and sanitization as well as large differences in the strength of virus emissions by infectious individuals can lead to highly variable number concentrations of airborne viruses in indoor environments. According to Leung *et al*. (2020) (*10*), the maximum numbers of viruses exhaled by certain individuals (*N*_v,30,ex_ ≈ 10^3^ to 10^5^) are several orders of magnitude higher than the mean values (*N*_v,30,ex_ ≈ 34 to 85). Such large variabilities are consistent with the distributions of viral load observed in respiratory tract fluids (*18, 19*), and we can fit the distribution of viral loads reported for SARS-CoV-2 by lognormal distribution functions with geometric standard deviations *σ* around ~1 to 2 (Sect S2).

The variabilities of viral loads, exhaled virus numbers, airborne virus concentrations, and inhaled virus numbers (*N*_v_) are key parameters in the overall assessment of average infection risks and the average efficacy of preventive measures for a population where large numbers of individuals are exposed to different conditions. For example, consider a scenario with a population-average inhaled virus number *N*_v,pop_ = 100 and an infectious dose ID_v,50_ = 100. If all individuals were exposed to the same airborne virus concentration and inhaled the same number of viruses, both the individual infection probability (*P*_inf_) and the population-average infection probability (*P*_inf,pop_) would be ~50% as given by Eq. 1, and masks would reduce the infection probability as illustrated for the virus-limited regime in Fig. 1C. If, however, 1% of the population were exposed to a very high concentration and inhaled 10,000 viruses while 99% of the population were not exposed and would inhale no virus, the population-average infection probability would be ~1%. In this extreme case, wearing masks would have no or little impact (low efficacy), because the individuals exposed to a very high concentration would be in a virus-rich regime and would mostly be infected with and without mask (*P*_inf_ ≈ *P*_inf,mask_ ≈ 1), while the others would remain uninfected with and without mask (*P*_inf_ ≈ *P*_inf,mask_ ≈ 0). In practice, the distribution of virus exposure and of the number of viruses inhaled by individuals in a population, the population-average infection probability, and the population-average efficacy of masks are expected to range between the above extremes.

To estimate population-average infection probability curves of *P*_inf,pop_ vs. *N*_v,pop_, we adopted lognormal distributions of *N*_v_ for SARS-CoV-2 with geometric standard deviations (*σ)* in the range of ~1 to 2 based on recently reported distributions of the viral load of SARS-CoV-2 in respiratory fluids (*18, 19*) (Sect S2). As shown in Fig. 2C, the slopes of these population-average infection probability curves are less steep in the virus-limited regime, and the range of transition to the virus-rich regime is broader than in case of uniform exposure (*σ* = 0).

Figure 3 illustrates how the efficacies of surgical masks, N95/FFP2 masks, and N99/FFP3 masks vary under different conditions (Sect. S4). Figure 3A shows the nonlinear increase of infection probability for individuals wearing a mask (*P*_inf,mask_) plotted against the infection probability without mask (*P*_inf_). Figure 3B shows the corresponding mask efficacy, i.e. the percentage reduction of infection probability (*ΔP*_inf_*/P*_inf_ = (*P*_inf_ *-P*_inf,mask_*)/ P*_inf_), which decreases slowly with increasing *P*_inf_ in the virus-limited regime, exhibits a steep decrease upon transition into the virus-rich-regime as *P*_inf_ approaches unity, and goes to zero at *P*_inf_ = 1. Figures 3C and 3D show equivalent plots for the population-average infection probability (*P*_inf,mask,pop_) and mask efficacy (*ΔP*_inf,pop_*/P*_inf,pop_ = (*P*_inf,pop_ *-P*_inf,pop,mask_*)/ P*_inf,pop_) in a population where the virus exposure is lognormally distributed with a geometric standard deviation of *σ* = 1. In the virus limited regime at small infection probabilities (*P*_inf,pop_ < 1%), the population-average mask efficacies (Fig. 3D) are similar to the individual mask efficacies in case of uniform exposure (Fig. 3B). In the transition range to the virus-rich regime, however, i.e., around *P*_inf,pop_ ≈ 0.5 corresponding to *N*_v,pop_ ≈ ID_v,50_, the population-average mask efficacies are lower.

**Fig. 3.**
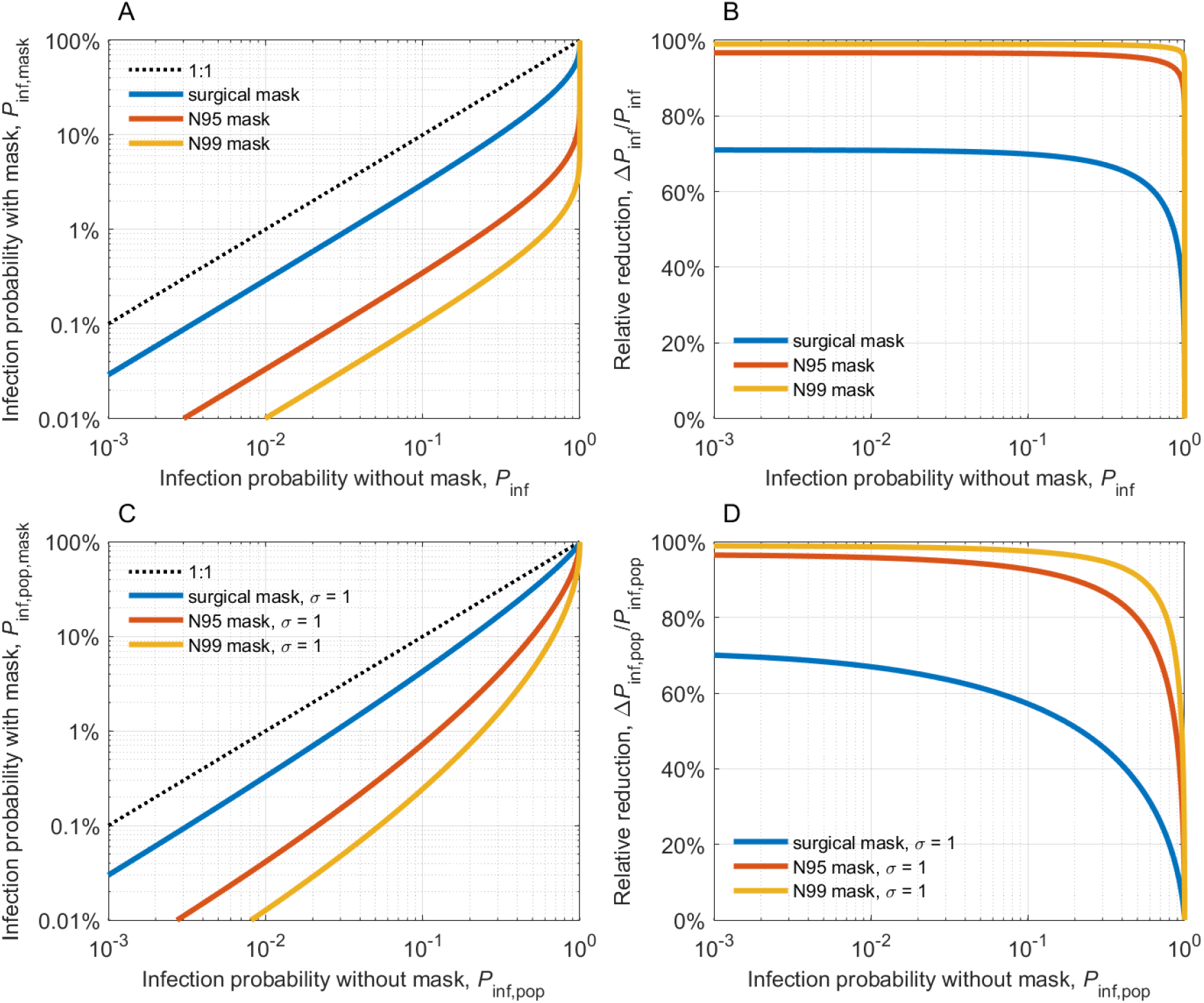
Reduction of COVID-19 airborne transmission by face masks. **(A)** Individual infection probability with mask (*P*_inf,mask_) plotted against individual infection probability without mask (*P*_inf_). **(B)** Individual mask efficacy, i.e., relative reduction of infection probability, *ΔP*_inf_*/P*_inf_, plotted against *P*_inf_. **(C)** Population-average infection probability with mask (*P*_inf,pop,mask_) plotted against population-average infection probability without mask (*P*_inf,pop_) assuming a lognormal distribution of virus exposure and *N*_v_ with a geometric standard deviation *σ* = 1. **(D)** Population-average mask efficacy, i.e., relative reduction of infection probability, *ΔP*_inf,pop_*/P*_inf,pop_, plotted against *P*_inf,pop_. The lines represent the results obtained for surgical masks (blue), N95/FFP2 masks (red), and N99/FFP3 masks (yellow), respectively.

The nonlinear dependence of mask efficacy on infection risk differs from the common assumption that the percentage change of infection probability due to mask use would be proportional to the percentage change of inhaled particle number. Under this assumption, wearing a mask would have the same impact on the transmission of a virus disease at any level of infection probability. Our analysis, however, shows that the efficacy of face masks depends strongly on the level of infection probability and virus abundance: masks are reducing the infection probability by as much as their filter efficiency for respiratory particles in the virus-limited regime, but much less in the virus-rich regime (Fig. 3). Accordingly, experimental investigations may find low mask efficacies when they are performed under virus-rich conditions, which may explain some of the apparently inconsistent results reported from randomized controlled trials (*5, 20, 21*). More importantly, the increasing effectiveness of mask use at low virus abundance implies synergistic effects of combining masks with other preventive measures that reduce the airborne virus concentration (e.g., ventilation and social distancing). The more measures are used, the more effective each measure will be in containing the virus transmission. For example, when both infectious and susceptible persons are wearing masks, the airborne virus concentration is reduced both at the source and at the receptor, thereby further improving mask efficacy through positive feedback. As the inhaled dose may also affect the severity of infections (*13*), masks can still be useful even if the reduced dose still leads to an infection.

For the transmission of SARS-CoV-2, a widespread prevalence of virus-limited conditions is indicated not only by the observational data and analyses presented above. It is also consistent with reported observations of the basic reproduction number of COVID-19 (*R*_0_ ≈ 2-7), the average duration of infectiousness (*d* ≈ 10 days), and the average daily number of human contacts (*c* ≈ 10-25) (*22–24*). According to the fundamental relation of these parameters (*R*_0_ = *P*_inf,pop_ ∙*c* ∙*d) (25*), we can estimate upper limit values in the range of ~0.8% to ~7% for the effective population-average infection probability of COVID-19 transmission, which is well below the infection probability expected for a virus-rich regime (*P*_inf_ ≈ 100%). This is consistent with the results of 172 observational studies across 16 countries and six continents which have shown a large reduction in the risk of infection by face mask use (*6*). Different regimes of abundance are relevant not only for the distinction of respiratory particles and viruses, but also for different types of viruses. For example, viruses with higher infectivity, i.e., with higher loads and rates of emission/exhalation, longer lifetime, or lower infectious dose, may result in a virus-rich regime and lead to higher basic reproduction numbers as observed for measles and other highly infectious diseases. Based on the scaling with ID_v,50_, the curves shown in Figs. 1 to 3 can easily be applied to assess the efficacy of masks and other preventive measures against new and more infectious mutants of SARS-CoV-2 such as B.1.1.7 (*26*).

Figure 4 shows the size distribution of particles emitted by different human respiratory activities (*27–29*). Our analysis of respiratory virus measurement data from ambient and exhaled air samples indicates that the “aerosol mode” (< 5 *µ*m) contains more viruses than the “droplet mode” (> 5 *µ*m), although the latter comprises a larger volume of liquid emitted from the respiratory tract (Table S2 and S7). This can be explained a higher viral load in the lower respiratory tract where the smaller aerosol particles are generated, and a lower viral load in the upper respiratory tract where the larger droplets are generated (*19, 30*). The enrichment of viruses in the aerosol mode can enhance their transmission, because smaller particles remain suspended for a longer time, leading to stronger accumulation and dispersion in the air, which may cause higher airborne virus concentrations, inhaled virus numbers, and infection risks – especially in densely occupied rooms with poor ventilation and long periods of exposure. Moreover, small aerosol particles have a higher penetration rate and higher probability to reach the lower respiratory tract (*31*), suggesting that they may cause more severe infections.

**Fig. 4.**
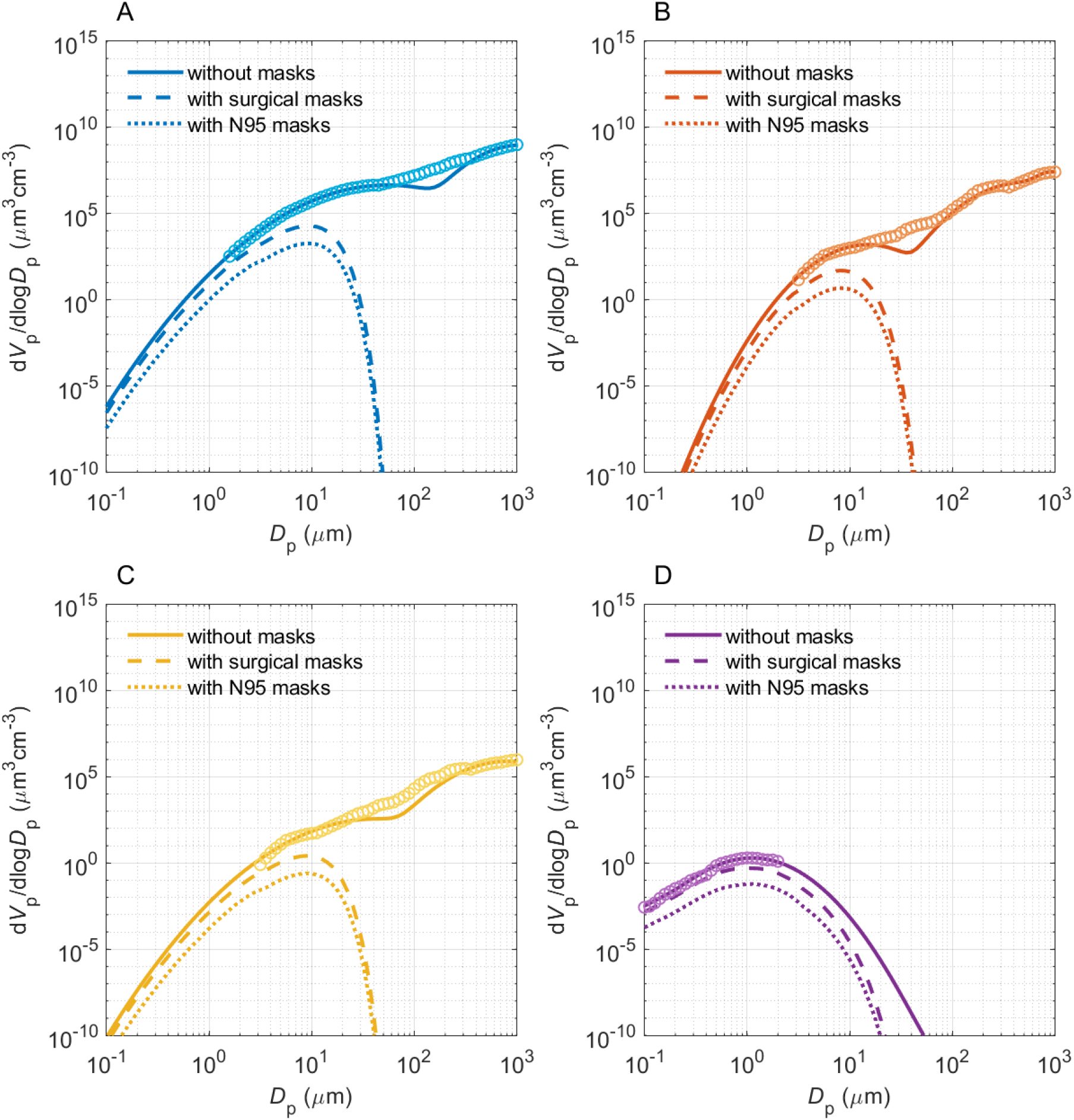
Volume size distributions of respiratory particles emitted during different respiratory activities with and without masks: sneezing (**A**), coughing (**B**), speaking (**C**), and breathing (**D**). The open circles are measurement data obtained without masks, and the solid lines are bimodal fits to the measurement data. The dashed and dotted lines are obtained by scaling with the filter efficiency curves of surgical masks and of N95/FFP2 masks, respectively (Sect. S4). The symbols *V*_p_ and *D*_p_ represent the particle volume and diameter, respectively.

The above analyses and discussions were focused on respiratory particles and droplets with diameters smaller than 100 *µ*m, which can remain suspended in the air over extended periods of time (traditional physical definition of aerosols). Because of rapid gravitational settling, respiratory droplets larger than 100 *µ*m are removed from the air within seconds, but they may still reach the upper respiratory tract of persons in close contact and cause infections by carrying large numbers of viruses in their very large liquid volume. For example, a single one-millimeter droplet may carry as many as ~50,000 viruses in case of a viral load of 10^8^ per milliliter respiratory fluid, which is realistic and higher than the estimated infectious dose for SARS-CoV-2 (*13*). Such super-large droplets, however, are very efficiently (~100%) removed even by simple masks (*32*), further emphasizing the importance and efficacy of face masks for preventing infections.

Our results have important implications for understanding and communicating preventive measures against the transmission of airborne viruses including SARS-CoV-2. When people see images or videos of millions of respiratory particles exhaled by talking or coughing, they may be afraid that simple masks with limited filtration efficiency (e.g., 30-70%) cannot really protect them from inhaling these particles. However, as only few respiratory particles contain viruses and most environments are in a virus-limited regime, wearing masks can indeed keep the number of inhaled viruses below the infectious dose and explain the observed efficacy of face masks in preventing the spread of COVID-19. However, the large variability of virus load may indeed lead to a virus-rich regime in certain indoor environments such as medical centers treating COVID-19 patients. Thus, medical staff should wear masks of high efficiency to keep the infection risk low. The nonlinear dependence of mask efficiency on the indoor virus concentration, i.e., the higher mask efficiency at lower virus abundance, also demonstrates the importance of combining masks with other preventive measures. Effective ventilation and social distancing will reduce ambient virus concentrations which further increase the mask efficiency in containing the virus transmission.

## Data Availability

All data is available in the main text or the supplementary materials.

(The following references cited only in SI)

## Acknowledgments

This study is supported by the Max Planck Society (MPG);

## Author contributions

Y.C. and H.S. designed and led the study. H.S., Y.C. and N.M. performed the research. U.P. and M.O.A. discussed the results. C.W., S.R. and P.W. commented on the manuscript. Y.C., H.S. and U.P. wrote the manuscript with inputs from N.M. and all coauthors;

## Data and materials availability

All data is available in the main text or the supplementary materials.

## Supplementary Materials

Supplementary Text S1 to S6

Figs. S1 to S9

Tables S1 to S7

References (*33-45*)

