## Supplementary Materials for "High efficacy of face masks explained by characteristic regimes of airborne SARS-CoV-2 virus abundance"

**This PDF file includes:**

Supplementary Text S1 to S6
Figs. S1 to S9
Tables S1 to S7

### Supplementary Text

#### S1. Scenarios in Leung *et al.* (2020)

Leung *et al.* (2020) reported an average of 5 to 17 coughs during 30-min exhaled breath collection for virus-infected participants (*10*). Taking the particle size distribution given in Fig. 4, we calculate that one person can emit a total number of $9.31\times10^{5}$ to $2.72\times10^{6}$ particles in a 30 min sampling period. Note that particles > 100 μm were not considered here, and the volume concentrations of particles in the "droplet" mode ($2.44\times10^{-4}$ mL, with $4.29\times10^{-5}$ to $2.45\times10^{-3}$ mL in 5% to 95% confidence level) overwhelm those in the "aerosol" mode ($7.68\times10^{-7}$ mL, with $3.37\times10^{-7}$ to $5.24\times10^{-6}$ mL in 5% to 95% confidence level).

#### S2. Virus concentration

##### S2.1 Virus concentration in exhalation samples of Leung *et al.* (2020)

Many samples in Leung *et al.* (2020) return a virus number below the detection limit (Fig. S1) (*10*). To reconstruct the whole distribution, we adopted an alternative approach, using the statistical distribution, i.e., percentage of positive cases, to calculate the virus number. Assuming that the virus number in the samples follows a Poisson distribution, the percentage of positive samples (containing > 2 viruses, i.e., Leung *et al.*, 2020 used $10^{0.3}$# as undetectable values in their statistical analysis) can be calculated with pre-assumed viral load in exhaled liquids. The Poisson distribution of virus number in emitted droplets is supported by early experiments, where the amount of bioaerosols or compounds delivered in particles is proportional to its concentration in the bulk fluid used to generate particles, and it is independent of investigated particulate type (fluorescent bead, bacteria or spore) (*33*).

For a set of sufficient samples, the positive rate (percentage of samples with virus number >2) is a function of the mathematical expectation of virus number per sample ($N_{\mathrm{v,sample,me}}$). The $N_{v\mathrm{,sample,me}}$ therefore can be retrieved by scanning a series of $N_{\mathrm{v,sample,me}}$ until the calculated positive rate agrees with the measurement. It should be noted that the viral load in exhaled liquids and the total exhaled liquid volume may be different among individuals, which must be considered in the calculation. Therefore, the Monte Carlo approach is used in this study. We assume that the statistical number distribution of SARS-CoV-2 in nasopharyngeal and nasal swab samples (Fig. S2, Jacot *et al.*, 2020 (*18*)) can represent the individual difference of viral load in exhaled liquids. The distributions can be fitted with multi-mode lognormal distribution with a low-abundance mode and a high-abundance mode. The dispersion of the high abundance mode ($\sigma$~ 1) is adopted in our calculation. In the experiment of Leung *et al.* (2020), the difference of sampled liquid volume stems from the individual difference of coughing times and volume concentration of exhaled droplets. The coughing time is assumed to follow a normal distribution with a $\sigma$ of 44.5. The exhaled droplet volume concentration is assumed to follow a lognormal distribution with a $\sigma$ of $\log_{10}(2)$. The experiment in Leung *et al.* (2020) is simulated with a series of viral loads. At each viral load, the experiment with the same sample number as in Leung *et al.* (2020) is repeated for $1\times10^{5}$ times to obtain a stable result. For each sample, the mathematical expectation of virus number is calculated based on randomly generated coughing time, exhaled aerosol/droplet volume concentration, and viral load. The "true virus number" in each sample is assumed to follow a Poisson distribution, and is randomly generated with its mathematical expectation equaling to the calculated value. Finally, for each pre-defined viral load, a distribution of positive rates can be obtained and fitted with a normal distribution function. When the calculated median positive rates become equal to the reported values in Leung *et al.* (2020), the viral load of coronavirus, influenza virus and rhinovirus in exhaled liquids are determined (Table S1), which is then used to calculate the distribution function and median of virus number per sample ($N_{\mathrm{v,sample}}$) (Fig. S3 and Table S2).

Given the volume of exhaled liquids in each sample ($V_{\mathrm{p}}$), the viral load in respiratory tract fluid, $C_{\mathrm{v,fluid}}$ (number of viruses per volume of respiratory tract fluid) can be calculated by:

$$C_{v,fluid} = \frac{N_{v,sample}}{V_p} \quad (1)$$

S2.2 Viral load in respiratory tract fluid of Jacot *et al.* (2020)

Jacot *et al.* (2020) presented a large 9-week dataset of viral load in nasopharyngeal and nasal swab samples (*18*). As shown in Fig. S2, the viral load apparently exhibited a multi-mode lognormal distribution (overall $\sigma \sim 2$), with one low-abundance mode around ~$1\times10^3$ to $1\times10^5$ copies $\mathrm{mL^{-1}}$, and the other high-abundance mode ~$1\times10^5$ to $1\times10^{10}$ copies $\mathrm{mL^{-1}}$. The high-abundance mode shows a negative skew lognormal distribution, probably due to the reduced viral load over time. To represent the individual difference of viral load in exhaled liquids, we took the variability of the high abundance mode with $\sigma \sim 1$. The low-abundance mode is not considered due to its much smaller contribution to the infection risk compared to the high-abundance mode.

**S3. Modelling of indoor airborne virus concentrations**

To compare the results of exhalation samples with indoor air samples, we performed model simulations for a scenario with patient density, space areas, and ventilation conditions emulating Fangcang Hospital in Wuhan:

- The total area of the ward is 500 $\mathrm{m^2}$ with a height of 10 m. The total number of patients is 200 (*14*).
- Each patient coughed an average of 34 times per hour, and the volume of each cough is 2 L; the breath volume is 8 L $\mathrm{min^{-1}}$. The size distributions of particles emitted during coughs and breath were taken from Fig. 4.
- All patients were wearing surgical masks with penetration rates given in Fig. S4A according to the guideline of Fangcang Hospital (https://edition.cnn.com/2020/02/22/asia/china-coronavirus-roundup-intl-hnk/index.html). We have also calculated the case when the patients did not wear any mask.
- Natural ventilation is assumed, and the loss rate of particles is calculated according to the function given in Fig. S5 (*34*).

The median viral load in exhaled samples were assumed the same as in Leung *et al.* (2020) (9) (Sect S2) and the variation between individual patients was assumed to follow a lognormal distribution with a $\sigma$ of 1. After being emitted, respiratory particles lose water and is dried to half of their initial sizes (*35*).

The indoor airborne virus concentration can be calculated with

$$C_v = 8 \cdot C_{v,aerosol} \cdot \int_0^{2.5\ \mu m} n(D_d) \cdot \frac{\pi \cdot D_d{}^3}{6} \cdot dlogD_d + 8 \cdot C_{v,droplet} \cdot \int_{2.5\ \mu m}^{\infty} n(D_d) \cdot \frac{\pi \cdot D_d{}^3}{6} \cdot dlogD_d \quad (2)$$

where, $C_{v,aerosol}$ and $C_{v,droplet}$ are the virus concentration in aerosol mode and droplet mode, respectively; $D_d$ is the particle dry diameter; $n(D_d)$ is the equilibrium indoor airborne particle number size distribution and can be determined by

$$\frac{dn(D_d)}{dt} = \frac{R_E(D_d)}{V} - \lambda(D_d) \cdot n(D_d) = 0 \quad (3)$$

where $R_E$ is the emission rate of particles by all patients; $V$ is the volume of the ward; and $\lambda$ is particle loss rate due to ventilation and deposition.

In the case when all patients were wearing surgical masks,

$$R_E(D_w) = R_{E0}(D_w) \cdot P_{mask}(D_w) \quad (4)$$

where, $R_{E0}$ is the emission rate of patients without wearing mask, $D_w$ is the wet diameter of exhaled droplets, $P_{mask}$ is size-resolved particle penetration rate of surgical masks. In this case, we assumed that exhaled liquid droplets only start to lose water after penetrating masks. In case no patients wearing masks, $R_E(D_w) = R_{E0}(D_w)$. Based on Eq. 3, the ambient particle number size distribution can be calculated as $n(D_d) = \frac{R_E(D_d)}{V \cdot \lambda(D_d)}$ when reaching equilibrium.

To account for the individual differences of viral load in exhaled particles, a Monte Carlo method is used to get the possible values of airborne virus concentration. The calculation is repeated for $1\times10^7$ times with randomly generated viral load, which follow a lognormal distribution with a $\sigma$ of 1. The calculated indoor airborne concentrations of coronavirus, influenza virus and rhinovirus are listed in Table S3.

Our calculation does not consider the lifetime of viruses (*36*). With a fixed virus emission rate, the airborne virus concentration is proportional to $\frac{1}{\lambda_v + \lambda_{dep} + k}$, where $\lambda_v$, $\lambda_{dep}$ and $k$ are loss rates due to ventilation, deposition and virus inactivation, respectively. The value of $k$ is similar as (or smaller than) $\lambda_v$ and $\lambda_{dep}$ (*37*). Therefore, ignoring virus loss due to inactivation ($k$) has a minor effect on the calculated airborne virus concentrations. The other caveat is that the particle loss rate ($\lambda_v$ + $\lambda_{dep}$) used here may differ from the real loss rate in Fangcang Hospital. According to the loss rate reviewed by Thatcher *et al.* (2002) (*38*), we may expect a maximum uncertainty of one order of magnitude in the calculated airborne virus concentrations, which will not change the regimes they belong to.

### S4. Penetration rate of masks and reduction of virus airborne transmission

The size-resolved particle penetration rate of surgical and N95/FFP2 masks (Fig. S4) is calculated based on the following literature and model calculation:

- Particle diameter < 800 nm: modified from Grinshpun *et al.* (2009) (*39*)
- Particle diameter > 800 nm & < 5μm: modified from Weber *et al.* (1993) (*40*)
- Particle diameter > 5μm: model calculation based on particle impaction with following parameters:

- Droplets velocity of 6.5 m $s^{-1}$, calculated based on the volume flow rate of 8 L $min^{-1}$ (typical breath flow rate of adults) and an air flow cross section as a circle with a diameter of 1 cm;
- Impact angle = 90 degree.

We assumed a filtration efficiency of 99% for N99/FFP3 masks. Regarding other simple masks, Drewnick *et al.* (2021) did a comprehensive evaluation of the filtration efficiency of household materials that can be used for homemade face masks and found huge differences of filtration efficiency between sample materials, spanning from <10% up to almost 100% (*41*).

The reduction of virus airborne transmission ($P_{inf}$ or $P_{inf,pop}$) by face masks in Fig. 3 is calculated from the change of $N_v$ based on the $P_{inf}$-$N_v$ or $P_{inf,pop}$-$N_v$ curves in Figs. 2. The change of $N_v$ is the sum of changes in both "aerosol mode" and "droplet mode". For each mode, the change of $N_v$ is assumed to be proportional to the change of volumes of respiratory particles by face masks.

### S5. Sample numbers and uncertainties

We found that the huge variability of the patient's exhaled virus concentration is an important reason for the contrast conclusions from experiments on efficacy of masks to prevent virus transmissions. This large variability requires a large number of samples to draw a robust conclusion. To illustrate the impact of the number of samples, a sensitivity experiment is conducted using a Monte Carlo approach: the virus number in samples of 30-min exhaled droplets above and below 5 µm is assumed to follow a lognormal distribution with median values as given in Table S2 and a $\sigma$ of 1. The sampling experiment is simulated with different sample numbers (2, 5, 10, 20, 50, 100, 200, 500 and 1000) and each experiment is repeated for $1\times10^4$ times. The standard deviation ($\sigma$) of the derived positive rates (percentage of samples with virus number > 2 (*10*)) is then calculated. Moreover, to see how the sample number influences the evaluation of the efficacy of masks, the virus number is calculated with a pre-assumed set of positive rates which follow a normal distribution with $\sigma$ shown in Fig. S6A. The frequency distributions of derived virus number in 30-min exhalation samples with and without masks at different sample numbers are given in Fig. S6B.

Figure S6A shows the variability of the positive rates under different number of samples. And Fig. S6B shows the frequency distributions of the calculated virus numbers under different sample numbers. When the number of samples is less than 10, the uncertainty of the observed positive rate is relatively large ($\sigma$ up to ~0.35), and the difference between the derived viral load in samples collected with and without mask use have a high chance to be indistinguishable (Fig. S6B). When the number of samples is ~ 100, the variability is small ($\sigma$ ~ 0.05), and the efficacy of masks become visible.

### S6. Effect of wearing masks

Early studies have calculated the effect of mask use on aerosol transmission and infection Risk of COVID-19 in different indoor environments (e.g., Lelieveld *et al.* 2020 (*42*)). Here, we evaluate the effect of wearing masks in controlling the SARS-CoV-2 virus transmission for a population.

As detailed below, wearing surgical masks may remove 82% of the SARS-CoV-2 virus. Because of the common existence of virus-limited regime, for simplicity, we assume that the percentage change of the virus transmission rate (i.e., the reproductive number) due to airborne transmissions is proportional to the percentage change of transmitted virus numbers. Given a basic reproduction number, $R_0$, of ~2.5 for COVID-19 (*23*), wearing a surgical mask can reduce it to ~ 0.46 and thus allow containing the virus. For N95/FFP2 masks, the reproductive number may even drop to 0.049. This degree of effect is apparently consistent with the real conditions (Fig. S7).

S6.1. Effect of wearing masks on reducing the reproduction number *R* of COVID-19

Wearing surgical or N95/FFP2 masks can reduce the emission rate of virus and further reduce the reproduction number *R* of COVID-19. Assuming that infectious individuals cough on average 20 times and speak for 10 minutes per hour, the volume emission rate of exhaled particles ($E_p$) can be calculated based on the size distributions shown in Fig. 4, and the number emission rate of virus ($E_v$) can be calculated with the viral load in Table S2. Table S4 shows the results for droplet size range of $D_w < 5\ \mu$m and $D_w < 20\ \mu$m. It can be seen that wearing surgical masks and N95/FFP2 masks can reduce the emission of virus by 81.7% and 98.0% ($D_w < 20\ \mu$m), respectively.

Assuming that the reproduction number *R* is proportional to the emission rate of viruses (*43*), the effect of wearing masks on *R* can be calculated. Assuming a basic reproduction number $R_0$ of 2.5, all infectious individuals wearing surgical mask and N95/FFP2 mask can reduce *R* to 0.46 and 0.049, respectively. It should be noted that only the mask removal of virus from the emitters is considered in the calculation. If all people wear masks, the number of viruses inhaled by healthy people will be further reduced, thereby further reducing *R*.

S6.2. The effect of wearing masks on the outbreak and popularity of COVID-19

To evaluate the effect of wearing masks on the dynamics of the COVID-19 outbreak, the infectious disease dynamics model (SEIR model) is employed to model the number of infections (*44*):

$$\begin{cases} \frac{dS_{pop}}{dt} = -\frac{\beta_t S_{pop} I_{pop}}{N_{pop}} \\ \frac{dE_{pop}}{dt} = \frac{\beta_t S_{pop} I_{pop}}{N_{pop}} - \sigma_i E_{pop} \\ \frac{dI_{pop}}{dt} = \sigma_i E_{pop} - \gamma_r I_{pop} \\ \frac{dR_{pop}}{dt} = \gamma_r I_{pop} \\ \beta_t = R_0 \gamma_r \end{cases} \tag{5}$$

where $N_{pop}$ is total population, $S_{pop}$ is the susceptible population, $E_{pop}$ is the exposed population, $I_{pop}$ is infectious population, $R_{pop}$ is recovered or dead population, $\beta_t$ is the transmission rate, $\sigma_i$ is the infection rate, $\gamma_r$ is the recovery rate, and $R_0$ is the basic reproduction number. Zhang *et al.* (2020) investigated the effect of limiting social contact patterns on the reproduction number of COVID-19 in Wuhan, China (*23*). We also select Wuhan as the target city, to compare the effects of wearing a mask and limiting social contact patterns reported in Zhang *et al.* (2020). The parameters in the SEIR model are assumed as follows (*23*, *45*):

- $N_{pop}$ = 11080000;
- $\gamma_r$ = 0.0556;

- $\sigma_i$ = 0.1923;
- $R_0$ = 2.5;
- The first outbreak occurred on December 2, 2019: $E_{pop}$ = 3000 and $I_{pop}$ = 10.

Assuming that control measures start on January 24 and no control measures are implemented before January 23, the effects of the following control measures are evaluated with the SEIR model:

- Only school closure: R = 1.9 (*23*);
- Reduce personnel contact (city lockdown through nonpharmaceutical interventions, such as home isolation, close public facilities, etc.): R = 0.34 (*23*);
- Wearing surgical masks, no other measures: R = 0.46;
- Wearing N95/FFP2 masks, no other measures: R = 0.049.

Figure S7A shows the results of the model calculation. Table S5 shows the cumulative total number of infections and the percentage of total infections under the five scenarios. It can be seen that wearing a surgical/N95/FFP2 mask can reduce the total infection rate to below 1%, which is similar as limiting social contact patterns. As a sensitivity study, we also calculated the total infection number and infection rate based on different virus emission reduction rates of masks. Results are shown in Fig. S7B and Table S6.

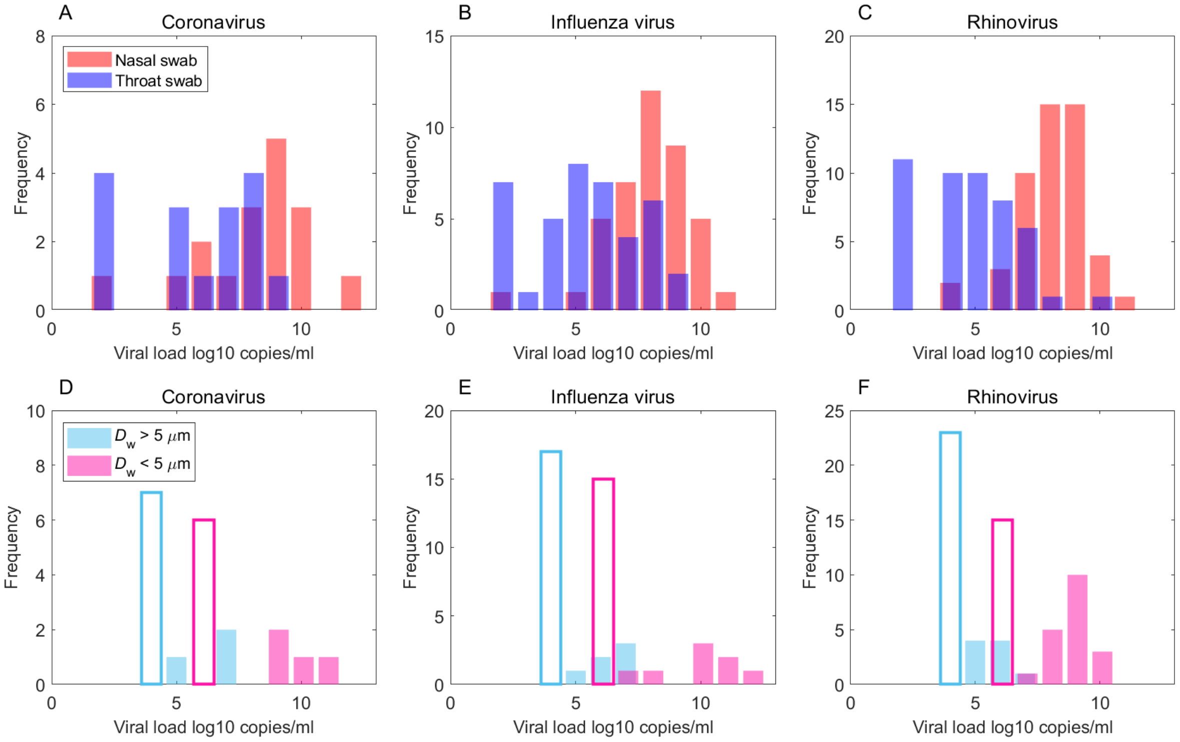


**Fig. S1. Frequency distributions of observed virus load in respiratory tract fluid. (A)**, **(B)** and **(C)** show the measured viral load in nasal and throat swabs for coronavirus, influenza virus and rhinovirus, respectively; **(D)**, **(E)** and **(F)** show the viral load calculated from virus number of exhalation samples (Eq. 1 in Sect S2). The unshaded bars represent samples with virus number below the detection limit (2 viruses, Leung *et al.* 2020 (*10*)).

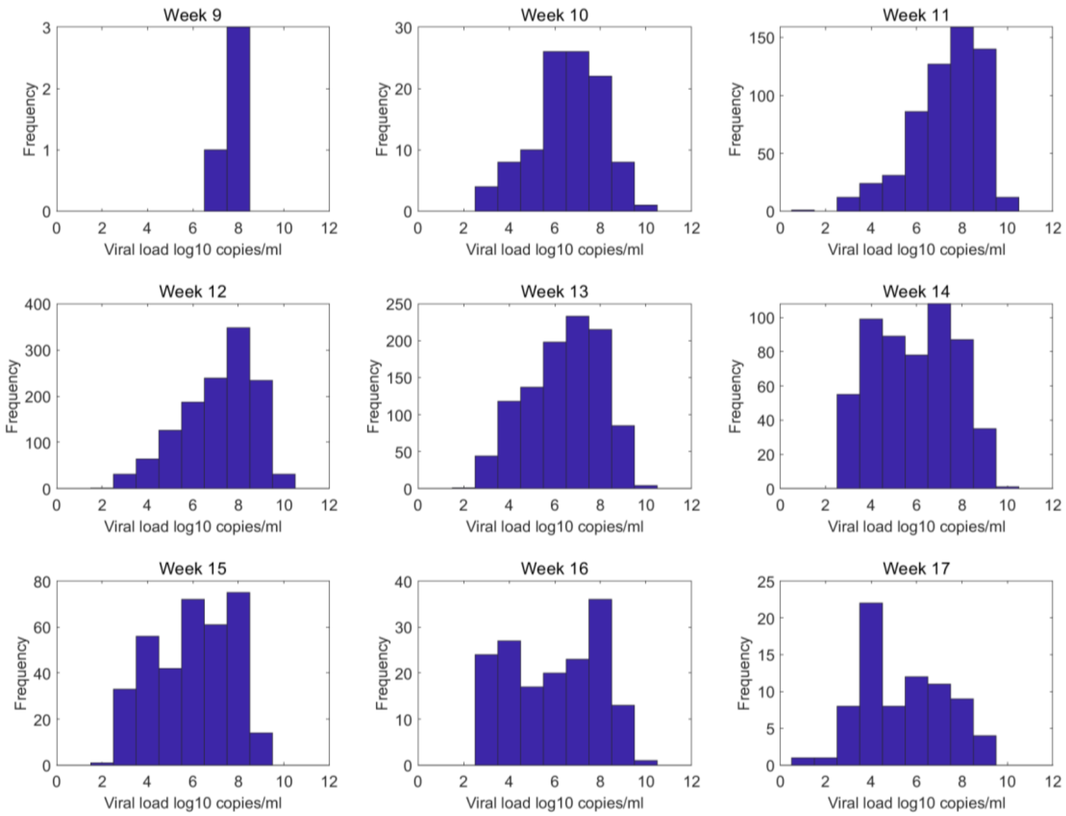


**Fig. S2. Frequency distribution of SARS-CoV-2 viral load in Jacot *et al.* (2020) (*18*).**

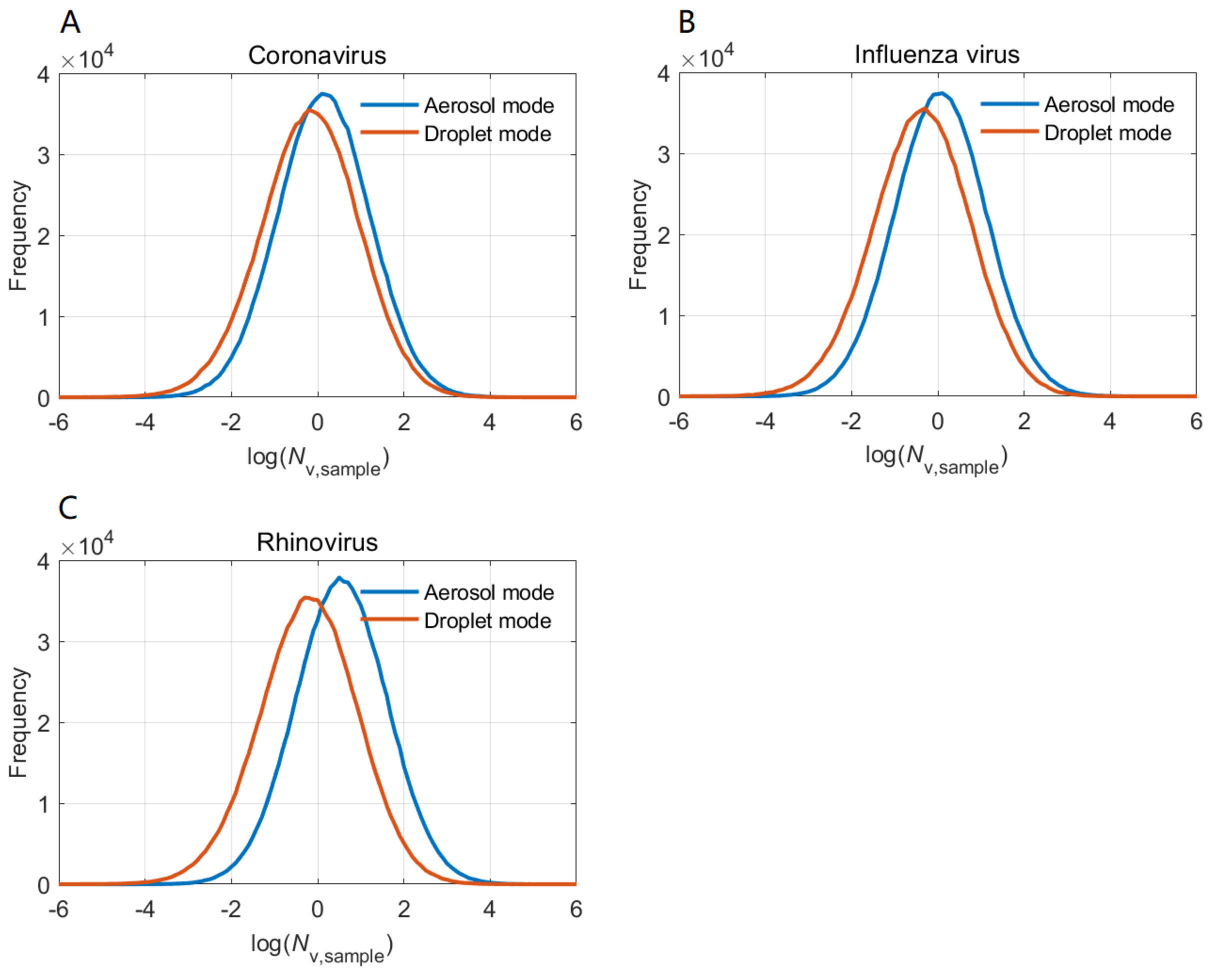


**Fig. S3. Frequency distributions of calculated virus number in 30-min exhalation air samples. (A)**, **(B)** and **(C)** show the distribution of coronavirus, influenza virus and rhinovirus, respectively. In each panel, the blue and red lines represent the virus number in aerosol mode and droplet mode, respectively.

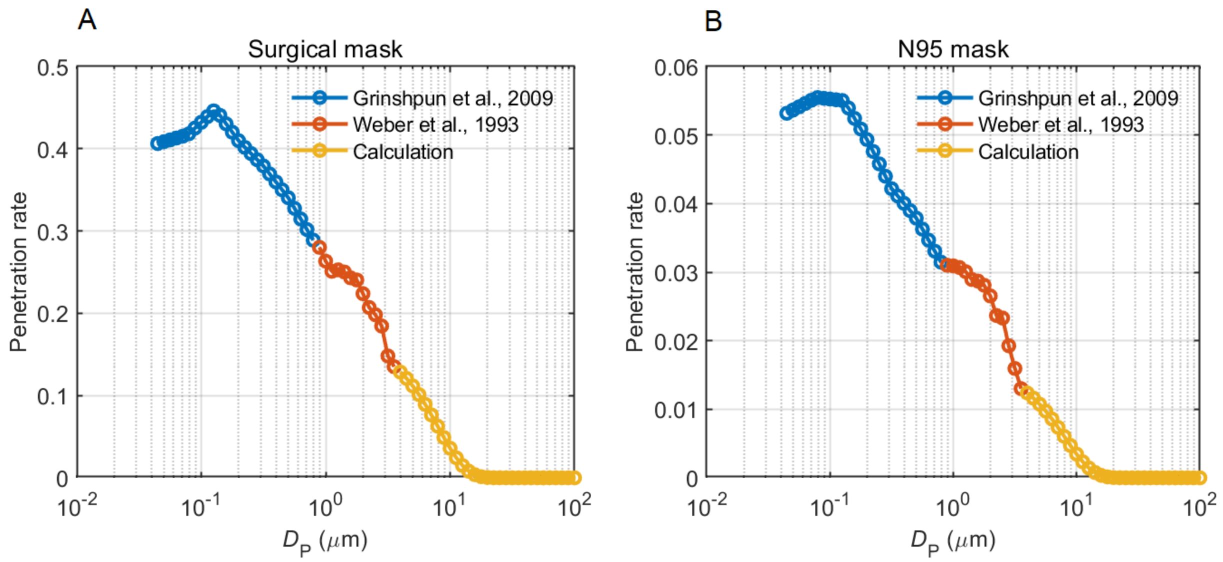


**Fig. S4. Particle penetration rate of a surgical mask (A) and a N95/FFP2 mask (B).** For the particle size range of ~50 nm to ~ 800 nm, the penetration rate (blue circle line) is modified from Grinshpun *et al.* (2009) (*39*). For particle size range of ~800 nm to ~ 3.5 $\mu$m, the penetration rate (red circle line) is modified from Weber *et al.* (1993) (*40*). For particle size above ~3.5 $\mu$m, the penetration rate (yellow circle line) is calculated based on particle impaction.

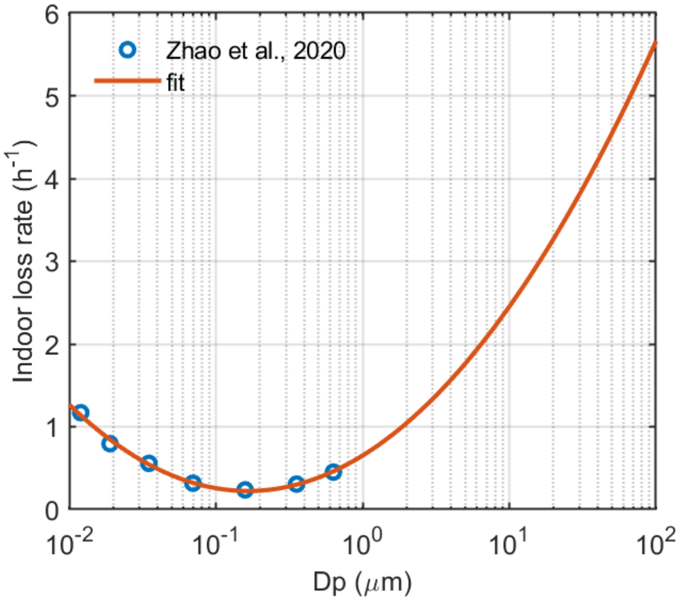


**Fig. S5. Size-resolved particle loss rate in indoor environment with natural ventilation.** The blue circles represent the measurement in Zhao *et al.* (2020) (*34*). The red line shows the fit result with $\lambda = 0.703 \cdot D_P^2 + 1.10 \cdot D_P + 0.651$.

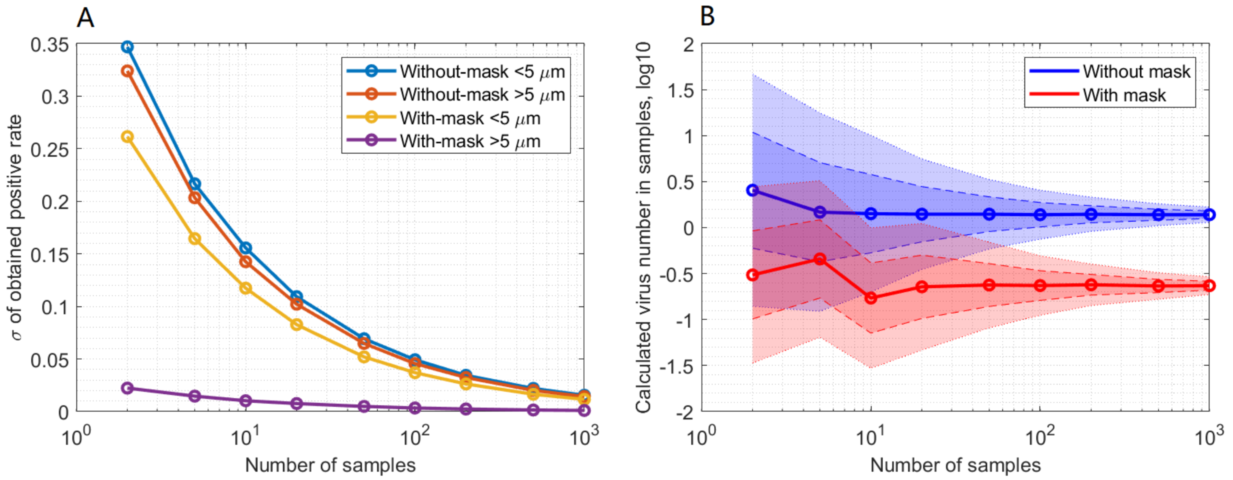


**Fig. S6. (A)** Standard deviation of positive rates derived based on different sample numbers. Four scenarios are tested: aerosol mode ($D_w < 5$ $\mu$m) samples of 30-min exhalation by patients without wearing surgical masks (blue circle line), droplet mode ($D_w > 5$ $\mu$m) samples of 30-min exhalation by patients without wearing surgical masks (red circle line), aerosol mode samples of 30-min exhalation by patients wearing surgical masks (yellow circle line), and droplet mode samples of 30-min exhalation by patients wearing surgical masks (purple circle line). The viral loads in aerosol and droplet mode particles are assumed to be the same as the coronavirus (Table S1). **(B)** Frequency distributions of derived viral load in 30-min exhalation samples at different sample numbers. The solid circle lines show the median viral load. Median $\pm\sigma$ and median$\pm 2\sigma$ are shown as dashed and dotted lines, respectively. The calculated viral load of coronavirus in aerosol mode (1.39# and 0.682#, Table S2) is adopted as the true viral load in the test. The positive rates of samples are assumed to follow normal distributions with $\sigma$ shown in panel **(A)**.

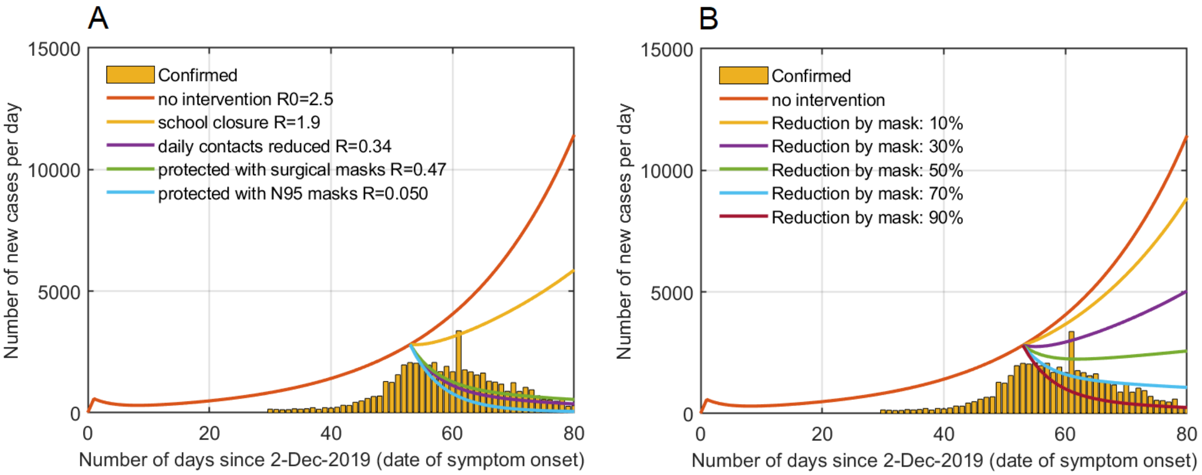


**Fig. S7. (A)** Reported daily new cases in Wuhan and simulated numbers based on different *R* and control measures. **(B)** Simulated daily new cases based on different virus emission reduction rates of masks. In panel **(A)** and **(B)**, the yellow bars represent the confirmed daily new cases in Wuhan and the colored lines show the simulated daily new cases by the SEIR model with different reproduction number *R*.

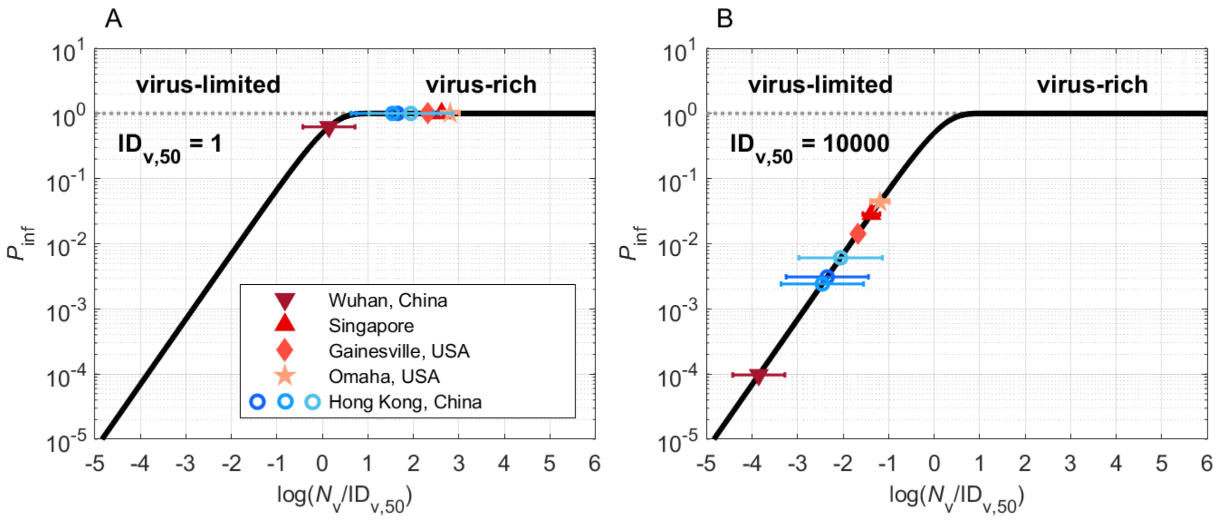


**Fig. S8.** The same as Fig. 2 except that the $ID_{v,50}$ is assumed to be 1 or 10000.

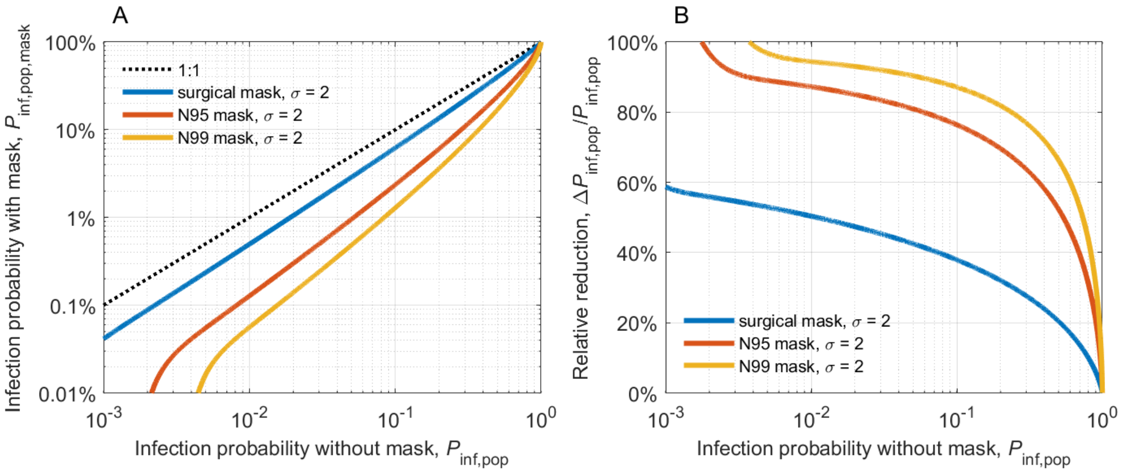


**Fig. S9. Reduced chance of COVID-19 transmission with masks.** The same as Fig. 3 except for using a $\sigma$ of ~ 2.

**Table S1. Viral load in exhaled liquids (# $mL^{-1}$).** The viral loads of coronavirus, influenza virus and rhinovirus in aerosol mode ($D_w < 5$ $\mu$m) and droplet mode ($D_w > 5$ $\mu$m) are retrieved based on the measured positive rates of 30-min exhalation samples (*10*). The individual differences of viral load and particle emission rate are considered in the calculation.

| | Coronavirus | Influenza virus | Rhinovirus |
|---|---|---|---|
| $D_w < 5$ $\mu$m * | $1.03\times10^6$ | $8.49\times10^5$ | $2.62\times10^6$ |
| $D_w > 5$ $\mu$m * | $1.55\times10^3$ | $9.69\times10^2$ | $1.42\times10^3$ |

* During the sampling of exhaled particles in Leung *et al.* (2020) (*10*), particles with size above and below 5 $\mu$m are separated very close to the mouth, thus the cut size (5 $\mu$m) of those two groups of particles is considered as wet diameter ($D_w$).

**Table S2. Virus number in the exhalation air samples (#).** The virus number in samples is calculated from the retrieved viral loads (Table S1) and total volume of exhaled particles during 30-min sampling.

| | Coronavirus | | Influenza virus | | Rhinovirus | |
|---|---|---|---|---|---|---|
| | Mean | Median | Mean | Median | Mean | Median |
| $D_w < 5$ $\mu$m | 27.3 | 1.39 | 23.2 | 1.15 | 69.5 | 3.55 |
| $D_w > 5$ $\mu$m | 17.7 | 0.682 | 10.9 | 0.429 | 15.3 | 0.627 |

**Table S3. Simulated indoor airborne virus concentration in Fangcang Hospital.** The indoor airborne concentrations of coronavirus, influenza virus and rhinovirus are simulated for two scenarios: virus emission by patients without wearing masks, and virus emission by patients wearing surgical masks. Median values and 5%, and 95% percentiles are given in the table.

| Scenarios | Coronavirus (# $m^{-3}$)<br>Mean (5%, 95%) | Influenza virus (# $m^{-3}$)<br>Mean (5%, 95%) | Rhinovirus (# $m^{-3}$)<br>Mean (5%, 95%) |
|---|---|---|---|
| Virus emission/exhalation by patients without wearing surgical masks | 5.49<br>(2.80, 10.3) | 3.78<br>(1.95, 7.04) | 7.88<br>(4.14, 14.7) |
| Virus emission/exhalation by patients wearing surgical masks | 0.400<br>(0.174, 0.837) | 0.327<br>(0.143, 0.675) | 1.01<br>(0.438, 2.10) |

**Table S4. Emission rate of droplet volume and virus number by infectious individuals.** The emission rates of droplets smaller than 5 $\mu$m ($D_w$ < 5 $\mu$m, $D_d$ < 2.5 $\mu$m) and smaller than 20 $\mu$m ($D_w$ < 20 $\mu$m, $D_d$ <10 $\mu$m) are given. Three scenarios, patients without wearing masks, patients wearing surgical masks, and patients wearing N95/FFP2 mask, are assumed in the calculation.

| Scenarios | $D_w$ < 5 $\mu$m ($D_d$ < 2.5 $\mu$m) | | $D_w$ < 20 $\mu$m ($D_d$ <10 $\mu$m) | |
|---|---|---|---|---|
| | $E_p$ (mL h$^{-1}$) | $E_v$ (# h$^{-1}$) | $E_p$ (mL h$^{-1}$) | $E_v$ (# h$^{-1}$) |
| No mask | $1.42\times10^{-6}$ | 20.3 | $4.16\times10^{-5}$ | 21.2 |
| Surgical mask | $2.69\times10^{-7}$ | 3.87 | $1.27\times10^{-6}$ | 3.89 |
| N95/FFP2 mask | $2.91\times10^{-8}$ | 0.418 | $1.26\times10^{-7}$ | 0.420 |

**Table S5. Total infection number and infection rate in Wuhan calculated based on different *R*.** The total infection number and infection rate are simulated with the SEIR model. The *R* for the control measures of school closure (*R*=1.9) and daily contacts reduced (*R*=0.34) are reported in Zhang *et al.* (2020) (*23*). And the *R* for the control measures of wearing surgical masks (*R*=0.46) and N95/FFP2 masks (*R*=0.049) are calculated assuming that *R* is proportional to the emission rate of virus-containing droplets.

| | *R* | Total infection number | Total infection rate |
|---|---|---|---|
| No intervention | 2.5 | $9.89\times10^6$ | 89.3% |
| School closure | 1.9 | $8.43\times10^6$ | 76.1% |
| Daily contacts reduced | 0.34 | $8.69\times10^4$ | 0.785% |
| Protected with surgical masks | 0.46 | $1.00\times10^5$ | 0.903% |
| Protected with N95/FFP2 masks | 0.049 | $6.81\times10^4$ | 0.614% |

**Table S6. Total infection number and infection rate in Wuhan calculated assuming different virus reduction rates of masks.** The total infection number and infection rate are simulated with the SEIR model. The reproduction number *R* is assumed to be proportional to the emission rate of virus-containing droplets.

| Reduction rate of mask | *R* | Total infection number | Total infection rate |
|---|---|---|---|
| 10% | 2.25 | 9.46E×$10^6$ | 85.4% |
| 30% | 1.75 | 7.91E×$10^6$ | 71.4% |
| 50% | 1.25 | 4.21E×$10^6$ | 38.0% |
| 70% | 0.75 | 1.82E×$10^5$ | 1.65% |
| 90% | 0.25 | 7.94E×$10^4$ | 0.717% |

**Table S7. Indoor airborne concentration ($C_v$) and 30-min inhaling number ($N_{v,30}$) of SARS-CoV-2 RNA copies in Fangcang Hospital.** The table is modified from Liu *et al.* (2020) (*14*). Room 1 and 2 are Protective Apparel Removal Room, and Room 3 is Medical Staff's Office. In the calculation of $N_{v,30}$, the total volume of inhaled air in 30 min is assumed to be 240 L.

| | Mode | Room 1 | Room 2 | Room 3 |
|---|---|---|---|---|
| $C_v$ (# $m^{-3}$) | Aerosol mode ($D_{amb}$ < 2.5 $\mu$m) * | 41 | 13 | 10 |
| | Droplet mode ($D_{amb}$ > 2.5 $\mu$m) * | 1 | 7 | 10 |
| $N_{v,30}$ (#) | Aerosol mode ($D_{amb}$ < 2.5 $\mu$m) * | 9.8 | 3.1 | 2.4 |
| | Droplet mode ($D_{amb}$ > 2.5 $\mu$m) * | 0.24 | 1.7 | 2.4 |

* In this study, the aerosol mode and droplet mode are defined as particles with wet diameter ($D_w$) smaller than 5 $\mu$m and lager than 5 $\mu$m, respectively. After being emitted, respiratory particles lose water and dry to ~ half of the initial particle size (*35*). Therefore, the boundary of these two modes for ambient particles is at ambient diameter ($D_{amb}$) of ~2.5 $\mu$m.
